# Knowledge and Practice of Emergency Contraception among Female Sex Workers: A Global Scoping Review

**DOI:** 10.1101/2025.04.01.25325048

**Authors:** M. Suchira S Suranga, K. Karunathilake, W. Indralal De Silva

## Abstract

**Introduction:** Female sex workers (FSWs) face a high risk of unintended pregnancies and abortion. Emergency contraception (EC) serves as a critical option for pregnancy prevention especially in the circumstances such as condom failure, stealthing, and sexual violence. Limited research has focused on the knowledge and practices of EC among FSWs. This scoping review aims to synthesize available evidence on the knowledge and use of EC among FSWs.

**Methods:** This review followed the Preferred Reporting Items for Systematic Reviews and Meta-Analyses extension for Scoping Reviews. A comprehensive search was conducted across four databases; Lens.org, Dimensions, PubMed, and Google Scholar. Inclusion criteria covered qualitative and quantitative journal articles published between 2000 and 2024 that examined EC knowledge and use among FSWs. Thematic analysis was performed and descriptive statistics were applied where relevant.

**Results:** The initial search yielded 735 studies, with 633 unique records after deduplication. Title and abstract screening shortlisted 34 articles for full-text review, of which 16 were excluded due to lack of relevance. An additional six studies were identified through reference screening, resulting in a final set of 24 studies. Findings revealed low awareness and usage of EC among FSWs despite a high prevalence of intentional and unintentional condom breakages, stealthing, sexual violence, and abortions. Median prevalence of life time use was 27.5%. Key barriers included misconceptions, cost, stigma, and lack of service availability.

**Conclusion:** FSWs’ experience indicated significant unmet needs for EC, yet research on this issue remains limited. Expanding access, addressing misinformation, and integrating EC into reproductive health services for FSWs are critical for reducing unintended pregnancies and associated health risks. Additionally, there is a pressing need for further research focusing on EC among FSWs in diverse settings to inform targeted interventions and policy development.

## 1. Introduction

Sex work is defined as the exchange of sexual services for money or its equivalent (1). It is a consensual sexual activity involving adults and young people of legal age, taking diverse forms across different countries and communities. The structure of sex work varies, ranging from informal, independent arrangements to highly organized systems (2).

Inconsistent condom use (3,4), limited adoption of non-barrier contraceptive methods including Long-Acting Reversible Contraceptives (5) and EC (6,7) along with a high partner exchange rate (4), restricted access to healthcare (8), and lack of bodily autonomy (9) contribute to significant health challenges among FSWs compared to the general female population (6). These risks are further exacerbated by high levels of sexual violence (10,11), coerced sexual acts, and the intentional or unintentional breakage and slippage of condoms by clients during intercourse (12,13,14). As a result, FSWs face numerous occupational hazards, including an increased risk of HIV and other sexually transmitted infections (STIs), unintended pregnancies, and both induced and spontaneous abortions (5,15). Table 1 presents data on the lifetime prevalence of induced abortion among FSWs in selected countries, highlighting the significant reproductive health burden faced by this population.

**Table 01:** Lifetime Prevalence of Induced Abortion among FSWs in Selected Countries.

|  | Country | Study | Sample Size (n) | Lifetime Prevalence (%) |
| --- | --- | --- | --- | --- |
| 01 | India | Shahmanesh et al, 2009 (16) | 325 | 28 |
| 02 | India | Wayal , et al., 2011 (17) | 326 | 26 |
| 03 | Bangladesh | Katz et al, 2015 (18) | 354 | 39 |
| 04 | Iran | Karamouzian, et al., 2016 (19) | 872 | 35 |
| 05 | Afghanistan | Todd et al, 2010 (20) | 520 | 27 |
| 06 | Colombia | Bautista, et al., 2008 (21) | 514 | 53 |
| 07 | Cambodia | Delvaux, et al., 2003 (22) | 632 | 25 |
| 08 | China | Lau, et al., 2007 (23) | 195 | 55 |
| 09 | China (Among adolescent FSWs) | Lim, et al., 2015 (24) | 310 | 93 |
| 10 | Benin | Sullivan, et al., 2020 (25) | 450 | 67 |
| 11 | England | Ward et al, 2000 (26) | 143 | 26 |
| 12 | Global Estimate | Mehrdad, K. et al., 2023 (15) | N/A | 38 |
| 13 | Countries where abortion is legal | Mehrdad, K. et al., 2023 (15) | N/A | 45 |
| 14 | Countries where abortion is illegal | Mehrdad, K. et al., 2023 (15) | N/A | 35 |

EC, also known as post-coital contraception, refers to methods used to prevent pregnancy within the first few days after unprotected or inadequately protected intercourse. It is intended for emergency use in cases of contraceptive failure or misuse such as missed pills or condom breakage as well as in instances of rape or coerced sex. However, EC does not provide protection against sexually transmitted infections (STIs), including HIV (27). For optimal effectiveness, EC should be used as soon as possible and within five days of intercourse. There are four main EC methods available globally: emergency contraceptive pills (ECPs) containing either levonorgestrel or ulipristal acetate, combined oral contraceptive pills (used in higher doses for EC), and copper-bearing intrauterine devices (IUDs), which are the most effective form of EC (28). World Health Organization (WHO) recommends that all women and girls at risk of unintended pregnancy have the right to access EC, and these methods should be routinely included within all national family planning programmes. Nonetheless, frequent and recurrent use of EC may carry potential risks for women with specific medical conditions. Additionally, it can lead to heightened side effects, such as menstrual irregularities, although it is worth noting that repeated use of EC is not associated with known health hazards (28).

While most research on the sexual and reproductive health (SRH) of FSWs has primarily focused on their vulnerability to sexually transmitted infections (STIs), particularly HIV, as well as unintended pregnancies and abortions (6), there has been limited attention on the use of non-barrier contraceptive methods, especially EC. Our comprehensive literature search did not identify any systematic review, narrative review, or scoping review that specifically examines the use of EC among FSWs. Addressing this research gap, the present scoping review aims to analyze the knowledge and practices related to EC among FSWs globally and identify the knowledge gap. The findings will provide valuable insights for future researchers conducting in-depth studies on EC use among FSWs and will support policymakers in developing more inclusive and integrated policies and programs to address unintended pregnancies and abortion within this population.

## 2. Methods

This scoping review on the knowledge and practices of FSWs regarding EC was conducted following the Preferred Reporting Items for Systematic Reviews and Meta-Analyses Extension for Scoping Reviews (PRISMA-ScR) guidelines (29). The research protocol was developed using PRISMA-ScR guidelines by the researchers and reviewed by 2 peer researchers who are familiar with Systematic Literature Reviews before finalized. A summary of the protocol is available as annexure 01. A comprehensive search was performed across four scientific databases and search engines; Lens.org, Dimensions, PubMed, and Google Scholar to identify relevant studies up to February 28, 2025. The search strategy incorporated Boolean operators to combine key concepts related to EC and FSWs, using the following terms: (“Sex Worker” OR “Prostitute*”) AND (“Emergency Contraceptive*” OR “Emergency Contraceptive Pill*” OR “Morning After Pill*”)*. Only peer-reviewed journal articles published between 2000 and 2024 were included in the review.

We adopted a broad definition of FSWs, including any female engaging in transactional sex, either full-time or part-time, as well as communities of women known to practice sex work, such as female entertainment workers in Bhutan and Cambodia (15). Both qualitative and quantitative studies published in peer-reviewed journals between 2000 and 2025 were included in the review (Figure 01).

**Figure 01:**
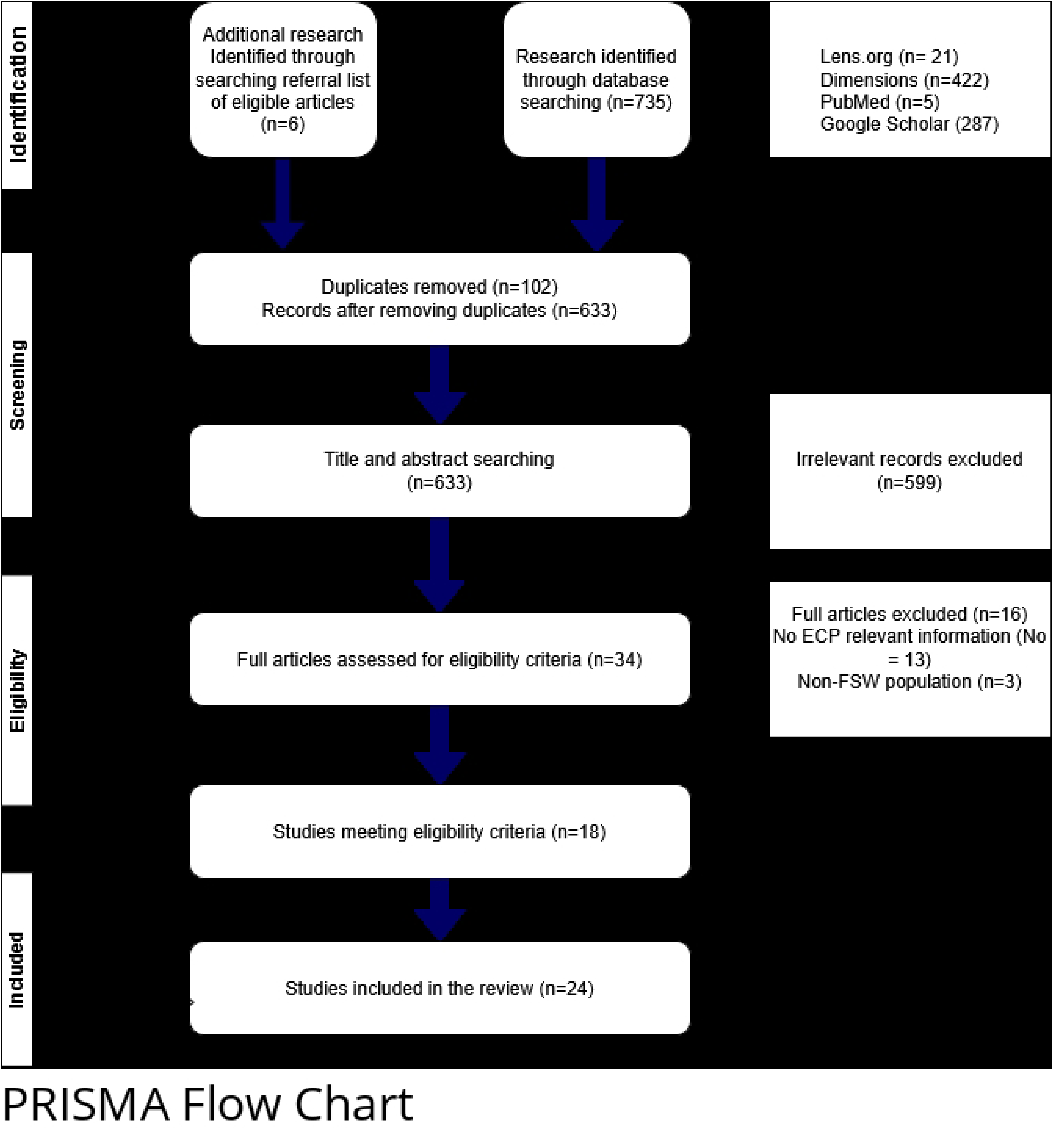
PRISM flow diagram for identification, screening and selection of studies

After removing duplicate records, the remaining unique studies underwent a two-stage screening process, conducted independently by two reviewers. In the first stage, the reviewers screened titles and abstracts for relevance, resolving any inconsistencies through discussion. In the second stage, the full texts of shortlisted articles were assessed against the eligibility criteria, again independently by both reviewers, with discrepancies resolved through discussion. Additionally, the reference lists of all included full-text articles were reviewed to identify any missed studies, leading to the inclusion of six additional studies. For non-English articles, Google Translate was used to facilitate comprehension during the screening process.

Data from the included studies were charted using a predefined extraction form developed in alignment with the review objectives. The form captured study characteristics, population details, and key findings related to knowledge and practices of emergency contraception among female sex workers. Two reviewers independently conducted data charting to reduce bias, resolving any discrepancies through discussion and consensus. The final dataset was reviewed collaboratively by the research team to ensure completeness and consistency before analysis. The data items and definitions included in the final data set is described in annexure 01. A formal critical appraisal of included studies was not conducted, in accordance with scoping review methodology; further details are provided in Annexure 01.

The final data set of all included articles was synthesized using a descriptive and thematic analysis approach. Quantitative data, such as prevalence figures, was summarized using basic descriptive statistics while qualitative findings was grouped and analyzed thematically under predefined categories such as knowledge of EC among FSWs, use of EC among FSWs, responses to condom failure and stealthing in relation to EC, the role of EC in cases of rape and sexual assault, factors associated with its use, and reasons for not using EC.

## 3. Results

The initial database search identified 735 potentially eligible studies, of which 633 were unique records after removing duplicates. Following the initial screening of titles and abstracts, 34 articles were selected for full-text review. During this stage, 16 articles were excluded, as three did not focus on FSWs, and 13 did not include information on EC. An additional six relevant studies were identified through reference list screening (Figure 01), resulting in a final set of 24 studies included in the review (Annexure 02).

### 3.1. Knowledge of EC among FSWs

Studies indicate that knowledge of ECs among FSWs is generally low, with only a few studies exploring their awareness and understanding of EC. While some FSWs are aware of EC, misconceptions regarding its correct usage, timing, and limitations are common across different regions.

A cross-sectional study conducted among 819 FSWs in Uganda found that 67.9% of participants had heard about EC (30). Similarly, a study among 466 FSWs in Côte d’Ivoire revealed that only one-third of all women in the study (n = 154) had knowledge of EC as a method to prevent pregnancy (31). In China, a study focusing on adolescent FSWs reported that around 64% had heard of EC (24). A multi-country cross-sectional study found that awareness of EC was significantly higher in high-income countries, with 74.8% of respondents demonstrating knowledge, compared to 96.1% in low and middle income countries (32).

Despite some level of awareness, studies highlight that FSWs often lack a comprehensive understanding of how EC should be used. A study conducted in Bahir-Dar town, North West Ethiopia, found that while 90% of the respondents had heard about EC, only 12% (n = 48) demonstrated a full understanding of its proper use (33). Another study from Ethiopia, which surveyed 346 FSWs, revealed that nearly half (52.9%, n = 183) had never even heard of EC. Among those who were aware, 38.6% (n = 63) mistakenly believed that EC could be used at any time, even after 120 hours following unprotected sex, illustrating widespread misconceptions about the method’s efficacy (34).

The knowledge of EC in specific contexts also varies. A study in Bhutan among 179 women working in the entertainment industry found that more than three-fourths (78.0%) of respondents knew that EC could be used after unprotected sex. However, significantly fewer recognized its applicability in cases of condom breakage (15.9%) or sexual assault (13.6%). While 71.2% of respondents could correctly identify the appropriate dosage, only 58.3% knew the correct timeframe for taking EC after sex. Encouragingly, 68.9% were aware that EC does not protect against HIV or other sexually transmitted infections (7).

Qualitative research further highlights the misconceptions surrounding EC use among FSWs. A study conducted in Kenya found that out of 30 respondents, only three knew about EC as an option to prevent pregnancy after condom failure. Some participants mistakenly believed that urinating immediately after condom failure would prevent pregnancy, demonstrating a critical gap in knowledge regarding effective post-coital contraceptive methods (12).

### 3.2. Use of EC among FSWs

As shown in Table 02, the current use of ECs among FSWs varies significantly across different countries. The lowest reported prevalence of current EC use was 3.1% in Uganda (30), while the highest was 21% in China (24). Similarly, the lifetime use of EC among FSWs ranged from 2.4% in India (35) to 50.7% in Nigeria (36), with a median of 27.5%, recorded in Swaziland (6). These findings align with a systematic literature review on abortion among FSWs, which reported that the lifetime prevalence of EC use among this population ranged from 2.4% in India to 38.1% in Kenya, with a median of 15.8% (15). Studies specifically examining the use of EC among adolescent and young FSWs (aged ≤20 years) are limited. A narrative review conducted on 2020 found that the proportion of FSWs below 20 years of age who have ever used EC was ranged from 5% (37) to 44% (24,26).

**Table 02:**
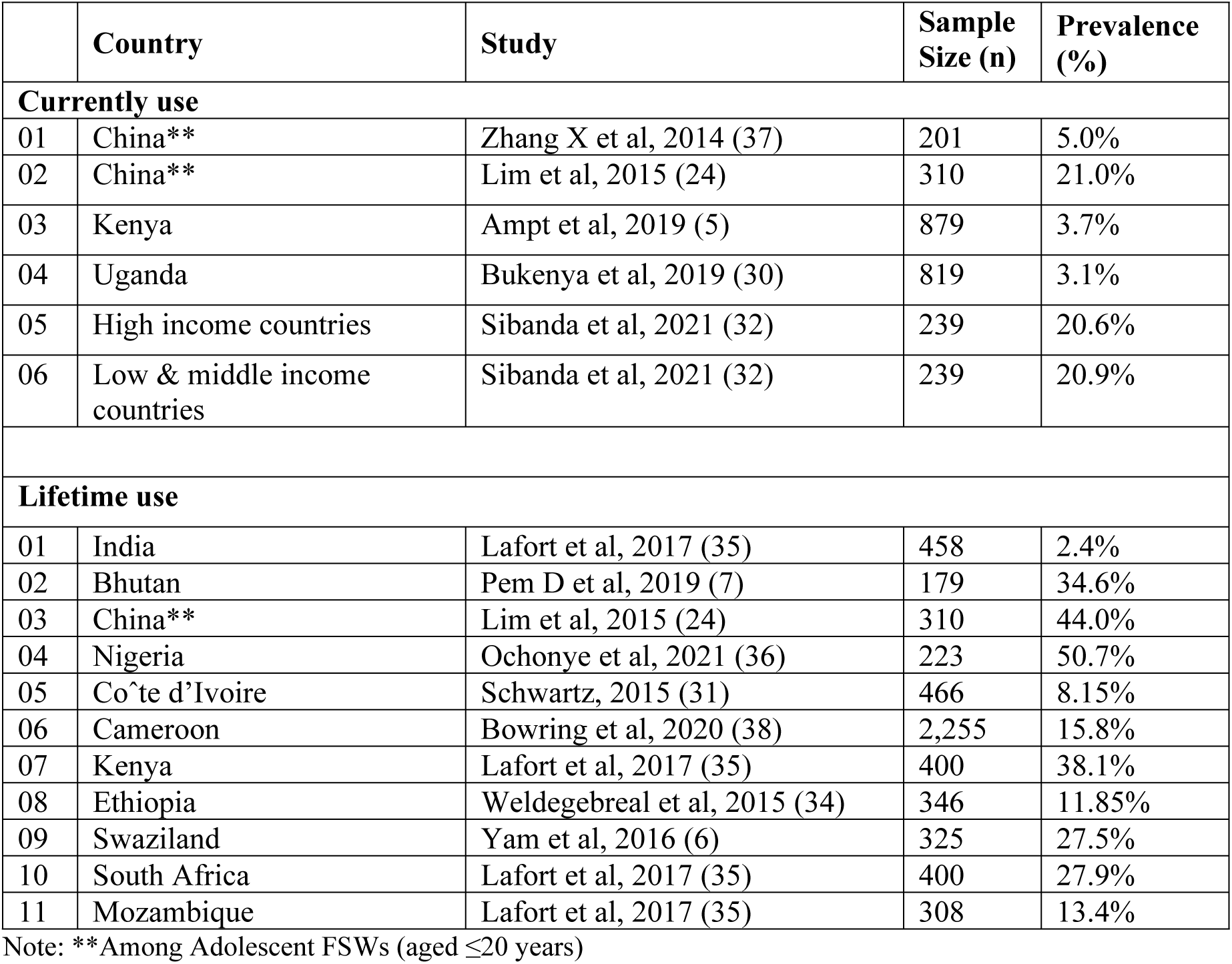
Prevalence of Current and Lifetime Use of EC among Female Sex Workers.

#### 3.2.1. Responses to Condom Failure and Stealthing: The Role of EC

Studies indicate that condom failure; whether through breakage, slippage, or intentional tampering by clients is a frequent occurrence among FSWs, posing significant risks for unintended pregnancies and sexually transmitted infections (14). The lifetime prevalence of condom failure among FSWs varies widely across different regions, ranging from 33% in China (39) to as high as 90% in Cape Town (13), with a median prevalence of 72% (Table 03). The responses of FSWs to condom failure vary significantly, often depending on their level of knowledge about EC and access to healthcare services. A qualitative study conducted in semi-urban Blantyre, Malawi, among 40 FSWs found that only 3 of the 18 women who experienced condom failure stopped sexual intercourse completely and sought medical care, including EC. Instead, most engaged in less effective post-exposure actions such as douching, urinating, or squatting in an attempt to prevent pregnancy (44).

**Table 03:** Prevalence of Condom Failure among Female Sex Workers in Selected Countries.

|  | Country | Study | Sample Size (n) | Prevalence (%) |
| --- | --- | --- | --- | --- |
| 01 | Ethiopia<sup>M</sup> | Wariso, et al., 2023 (40) | 6085 | 29.9% |
| 02 | Ethiopia<sup>M</sup> | Amogne, et al., 2019 (41) | 4,900 | 25.5% |
| 03 | Bangladesh<sup>M</sup> | Katz et al, 2015 (18) | 354 | 51.0% |
| 04 | China<sup>Q</sup> | Choi, et al., 2008 (42) | 200 | 36.2% |
| 05 | China<sup>LT</sup> | Qu, et al., 2002 (39) | 482 | 33.0% |
| 06 | China<sup>LT</sup> | Lau, et al., 2007 (23) | 195 | 86.2% |
| 07 | Greece<sup>LT</sup> | Apostolidou, et al., 2023 (43) | 71 | 63.5%** |
| 08 | Malawi<sup>LT</sup> | Twizelimana & Muula, 2020 (44) | 40 | 45.0% |
| 09 | Kenya<sup>LT</sup> | Masvawure, et al., 2018 (14) | 75 | 81.0% |
| 10 | Cape Town<sup>LT</sup> | Mukumbang, 2017 (13) | 100 | 90.0% |

**Note** : M=Prevalence during past 30 days, Q=Prevalence during past 3 months, LT = Lifetime Prevalence, \*\*= Prevalence of stealthing, the nonconsensual condom removal in sex

A quantitative study conducted among 100 FSWs in Cape Town found that while 90% had experienced condom breakage and 85% had experienced condom slippage at some point in their lives, only 8% reported using EC afterward. Instead, 53% took no action, while others engaged in ineffective protective behaviors such as applying vaginal spermicidal foam (3%) or washing immediately (5%). Notably, 3% reported consuming alcohol or drugs to cope with the incident (13). A separate qualitative study among 30 FSWs in Kenya confirmed that condom failure was a frequent occurrence. Many women attempted to mitigate risk by ensuring correct condom use, providing their own condoms, and applying them themselves. In the aftermath of condom failure, some women sought medical attention, particularly for sexually transmitted infections, but very few mentioned EC as an option (12).

Intentional condom tampering by clients and nonconsensual condom removal (stealthing) pose serious risks to the health and safety of FSWs, further increasing their vulnerability to unintended pregnancies and sexually transmitted infections (12,14,43). A qualitative study conducted in Kenya found that 81% of participants had experienced condom breakage or slippage during commercial sex. The study categorized condom failure into two main types: unintentional failures caused by factors such as inebriation, forceful thrusting, or incorrect condom use, and intentional failures attributed to deliberate actions by clients, such as covertly removing or puncturing the condom. Similarly, research in Malawi highlighted that a significant number of FSWs had encountered clients who deliberately removed condoms often under coercive circumstances (44). A study conducted in Greece among 71 FSWs reported that 63.5% had experienced stealthing, a significantly high prevalence that highlights the widespread nature of this violation (43). Responses to condom failure varied widely, with some women replacing the condom and resuming sex, while others resorted to ineffective protective measures like washing. Only a few reported using EC following condom failure (14).

#### 3.2.2. Rape, Sexual Assault, and the Role of EC

Sexual violence is a major public health concern and a violation of human rights (45). FSWs are at an increased risk of rape and sexual assault, often due to criminalization of sex work, lack of legal protection, and unsafe working environments. Many FSWs operate in isolated locations, such as dark alleys, abandoned buildings, or unfamiliar hotels, increasing their vulnerability to violent clients (10). Studies from different countries report high rates of rape and sexual assault among FSWs, with prevalence ranging from 10.9% to 57.9%, depending on the region and timeframe considered (Table 04). FSWs who reported forced sex were 2.69 times more likely to experience an unwanted pregnancy compared to those who had not been raped (11).

**Table 04:** Prevalence of rape or sexual assaults among Female Sex Workers in Selected Countries.

|  | Country | Study | Sample Size (n) | Prevalence (%) |
| --- | --- | --- | --- | --- |
| 01 | Ethiopia<sup>Y</sup> | Misganaw & Worku, 2013 (33). | 395 | 11.4% |
| 02 | India<sup>Y</sup> | Patel, et al., 2016 (46) | 2,400 | 24.0% |
| 03 | South Africa<sup>Y</sup> | Jewkes, et al., 2021 (47) | 3005 | 57.9% |
| 04 | Uganda<sup>2Y</sup> | Bukenya, et al., 2019 (30). | 819 | 21.0% |
| 05 | Canada<sup>2Y</sup> | Shannon, et al., 2009 (48) | 237 | 44.0% |
| 06 | Ethiopia<sup>SW</sup> | Amogne, et al., 2019 (41) | 4,900 | 15.2% |
| 07 | Sri Lanka<sup>LT</sup> | Manathunge, et al., 2020 (4) | 442 | 10.9% |
| 08 | Ethiopia<sup>LT</sup> | Misganaw & Worku, 2013 (33). | 395 | 24.3% |
| 09 | Gambia<sup>LT</sup> | Sherwood, et al., 2015 (11) | 251 | 29.0% |
| 10 | Uganda<sup>LT</sup> | Schwitters, et al., 2015 (49) | 1467 | 49.0% |

**Note** : LT = Lifetime, SW= since they start sex work, 2Y= during the last two years, Y= during the last year,

Despite the high prevalence of sexual violence among FSWs, research on the use of EC following rape remains scarce. The limited studies available suggest that only a small proportion of survivors access EC after experiencing sexual violence. For instance, a study of 395 street-based FSWs in Ethiopia found that 24.3% had experienced rape in their lifetime, and 11.4% had been raped in the past year. None of the victims used condoms during the assault, and only 4 (4.2%) used EC post-rape (33). Similarly in Uganda, a study among 819 FSWs found that 172 (21.0%) had been raped in the past two years, but only 70 (40.7%) accessed EC after the assault (30).

However, a qualitative study in Nepal recognized EC as a lifesaving intervention for FSWs who experience sexual violence. Supervisors acknowledged that post-rape EC was crucial but often underutilized due to barriers in access (10). Failure to use EC after unprotected forced sex increases the risk of unintended pregnancies among FSWs, particularly those not using modern contraceptive methods (30).

#### 3.2.3. Factors Associated with Use of EC

Several factors have been identified as influencing the use of EC among FSWs. A cross-sectional study conducted among 325 FSWs in Swaziland found that education level was significantly associated with EC use. Women with secondary education were nearly four times as likely to have used EC compared to those with primary education or less. Additionally, age was a strong predictor of EC use, with younger women being more likely to use EC. Women aged 25 years or older had lower odds of using EC compared to their younger counterparts. Marital status also played a key role in EC use. Single FSWs were significantly more likely to have used EC, with 34.9% of single women reporting EC use compared to 16.7% of ever-married women (p = 0.029). In fact, single women had more than three times the odds of using EC compared to those who had ever married. Income was another factor associated with EC use. Higher-income FSWs were more likely to use EC, particularly when the partner exchange rate increased (p = 0.006). Further, condom use and condom failure were strongly linked to EC use. Women who had not always used condoms in the past month were significantly more likely to have used EC (38.1% vs. 15.8%; p < 0.001). Similarly, those who had experienced condom failure in the past month were more likely to have used EC (40.7% compared to 23.1%; p = 0.001) (6).

A national-level study conducted in Cameroon involving 2,255 FSWs also identified that a history of termination of pregnancy was associated with a higher likelihood of EC use. FSWs who had previously terminated a pregnancy were 34% more likely to have used EC compared to those who had not. Additionally, the study found that FSWs who had ever used EC were 2.7 times more likely to use non-barrier contraceptives (such as hormonal methods, intrauterine devices (IUDs), or sterilization) compared to those who had never used EC (38).

#### 3.2.4. Reasons for Not Using EC

Despite the potential of EC in preventing unintended pregnancies, many FSWs are reluctant to use it. A study conducted among 395 street-based FSWs in Ethiopia found that the majority (95.8%) did not use EC, citing a variety of reasons. The most common reason was reluctance, reported by 41.8% of participants, followed by fear of side effects (19.8%). Other barriers included difficulty in identifying clinics that offered appropriate services (12%), lack of money (10.9%), lack of awareness about EC (10.9%), and fear of stigma (7.8%) (33).

A similar study in Bhutan revealed that 79.9% of respondents had concerns about the potential harm of EC to their bodies. The most prevalent concerns included its effect on future pregnancy (76.1%), side effects (70.2%), and an increased risk of HIV/STIs (69.4%) (7).

## 4. Discussion

Despite the critical importance of EC for FSWs, only a few studies have examined their knowledge, attitudes, and practices (KAP) regarding EC globally (6). However, the need for EC among FSWs is acute due to high rates of sexual violence, condom failures, unintended pregnancies, and abortions experienced by this population. Research has shown that EC use can significantly reduce unintended pregnancies and abortions (50). However, where reported, EC use among FSWs remains low in many settings (51). Those from lower socioeconomic backgrounds, who are at the highest risk of unintended pregnancies, often face significant barriers to EC access, including cost and lack of availability (6).

The strong association between condom failure, stealthing, and EC use suggests that condom breakage and slippage are key drivers of EC use among FSWs (12,14,44). These findings underscore the need for programmatic interventions that address condom failure, including ensuring access to EC as a backup method (6). Findings from this study confirm the high prevalence of sexual violence and forced sexual activity among FSWs (33,10,11). These results highlight the need for an integrated response to sexual violence, including immediate access to EC for survivors of forced sex (11). Strengthening post-rape care services and ensuring that EC is part of standard care protocols for sexual violence survivors could help mitigate the risk of unintended pregnancies.

A comparative study among FSWs in Nigeria found that those who received targeted interventions were 2.22 times more likely to use EC than their baseline counterparts (36). This study demonstrated the effectiveness of educational and intervention programs in increasing EC uptake. Similar programs should be adapted and implemented in other settings to improve access and awareness among FSWs. However, it must be acknowledged that as FSWs are a marginalized and hard-to-reach population, reproductive health interventions, particularly those aimed at increasing EC access must be tailored to their unique needs (34). Social and behavior change interventions, delivered by peer educators or healthcare providers, could address misinformation, fears, and limited knowledge about EC. Additionally, efforts to promote dual-method contraception should emphasize the use of long-acting reversible contraception (LARC) alongside EC as a backup option (52).

Healthcare providers play a crucial role in increasing EC awareness and uptake. Training programs should focus on improving providers’ counseling and communication skills, ensuring that they discuss the full range of contraceptive options, including EC as part of comprehensive reproductive health counseling (52). This is particularly important in cases of forced sexual intercourse (33) and condom failure (14,44).

Given the high risk of unintended pregnancies and induced abortions in this population, ensuring access to EC is essential, particularly in the context of inconsistent condom use, frequent condom failure, and sexual violence. However, EC has a relatively high failure rate compared to other modern contraceptive methods and should not be relied upon as a primary method of contraception (27). Over-reliance on EC instead of more effective long-term contraceptive methods remains a concern in many populations (50) including FSWs (Lim et al., 2015).

Through a comprehensive and systematic literature review, this study identified a significant gap in research on the KAP of EC among FSWs globally. Very few studies have specifically focused on EC use in this population, with most research on contraception uptake overlooking EC. Future research should prioritize a more in-depth examination of EC knowledge, attitudes, and practices among FSWs in diverse settings to develop effective, context-specific interventions.

Legal frameworks governing sex work play a critical role in shaping female sex workers’ (FSWs) access to and use of EC. In fully criminalized settings, such as Swaziland and Uganda, FSWs face significant barriers including fear of arrest, confiscation of condoms, and provider stigma which lead to low EC utilization despite high rates of unprotected sex, condom failure, and sexual violence (6,30,53). Partial criminalization models, like the Nordic approach, similarly displace sex workers into unsafe conditions, perpetuate stigma, and discourage healthcare access, resulting in limited EC use and poor sexual health outcomes (53). In legalized systems, such as in parts of Australia and Europe, registered FSWs may benefit from structured healthcare access and improved EC availability, but those outside the legal framework due to licensing barriers or undocumented status remain excluded and underserved (54,53). In contrast, decriminalized environments like New Zealand and New South Wales demonstrate significantly better outcomes, including improved EC access, stronger peer-led outreach, reduced stigma, and lower rates of unintended pregnancy, indicating that decriminalization fosters an enabling environment for reproductive health (54).

This scoping review has several limitations. One significant limitation is the lack of research specifically focused on EC use among FSWs, which resulted in a limited number of studies available for review. Furthermore, the included studies varied in design, methodology, and geographical context, which may have introduced inconsistencies in the findings and hindered the ability to draw broad generalizations. Additionally, some studies combined EC use with other contraceptive methods, making it difficult to isolate the specific knowledge and practices related to EC. Finally, due to the limited research in this area, there is a significant gap in understanding the contextual factors that influence EC use among FSWs, particularly in low- and middle-income countries.

## 5. Conclusion

This study underscores low level of awareness and use of ECP with the median prevalence of life time use of 27.5% among FSWs. This indicates the critical need to enhance access to EC among FSWs due to the high prevalence of unintended pregnancies, induced abortion, and sexual violence, including rape and assault. Intentional and unintentional condom breakage, slippage, and stealthing by the clients further increase the risk of unintended pregnancies, yet knowledge and use of EC remain low. Limited research on EC use among FSWs at the global level hinders effective policy and program development. Addressing these gaps through targeted education, improved access to EC, and stronger protections against sexual violence is essential for safeguarding the reproductive health and rights of FSWs.

## Data Availability

The data underlying the results presented in the study are available from the relevant research articles included in the list of references.

## External Funding

Authors declare that no external funding.

## Ethical Approval

This study was based solely on secondary data and a review of existing literature, without the involvement of human or animal subjects; therefore, ethical approval was not required for this phase. However, ethical clearance for the broader research project, which includes a subsequent stage involving human participants, has been obtained from the Ethics Review Committee of the University of Kelaniya, Sri Lanka (UoK/ERC/MDS/2024/038).

## Acknowledgements

The authors express their sincere gratitude to the following individuals for their invaluable contributions to this study: Dr. KGS Priyashantha, Senior Lecture, University of Peradeniya, Dr. Ruchitha Perera, Executive Director of The Family Planning Association, for his expertise and guidance. Mr. Duminda Rajakaruna, Mr. Janaranga Dewasurendra, and Mr. Amal Bandara for their dedicated support and assistance. Dr. Lakshman Senanayake for his valuable insights and contributions to the research. These individuals played a crucial role in enriching the quality and depth of the study, and their collaboration is deeply appreciated.

## Use of Artificial Intelligence Assisted Technologies

During the preparation of this work the authors used ChatGPT in order to improve the language and readability. After using this tool/service, the authors reviewed and edited the content as needed and takes full responsibility for the content of the publication.

## Author contributions

SS played a key role in the conceptualization, data curation, analysis, and drafting of the manuscript. KK and IDS contributed significantly to the screening and selection of articles, provided ongoing supervision throughout the process, and were actively involved in reviewing and finalizing the manuscript.

## Annexure 01: Review Protocol Summary

|  |  |
| --- | --- |
| **Study Title:** | Knowledge and Practice of Emergency Contraception among Female Sex Workers: A Global Scoping Review |
| **Authors:** | M. Suchira S Suranga<sup>1*</sup>, K. Karunathilake<sup>2</sup>, W. Indralal De Silva<sup>3</sup><br><sup>1</sup>The Family Planning Association of Sri Lanka, Sri Lanka; <sup>2</sup>University of Kelaniya, Sri Lanka. <sup>3</sup>University of Colombo, Sri Lanka. |
| **Study Type** | Scoping Review |
| **Objective** | The objective of this scoping review is to explore and synthesize existing global evidence on the knowledge and practices related to emergency contraception among female sex workers, with the aim of identifying key patterns, gaps, and implications for future research and public health interventions. |
| **Article selection method** | PRISMA-ScR guidelines |
| **Search strings** | Sex Workers, Prostitute Emergency Contraceptive, Emergency Contraceptives, Emergency Contraceptive Pills, Morning After Pills |
| **Inclusion criteria** | 1. Year range: 2000 February 2025<br>2. Subject area: All<br>3. Language of article: Any<br>4. Keywords: Sex Workers, Female Sex Workers, Women Sex Workers, FSW, WSW, Emergency Contraception, Emergency Contraceptives, Emergency Contraceptive Pills, EC, ECP, Morning after Pills,<br>5. Source type: Academic Journals<br>6. Type of Study: Empirical and meta analysis<br>7. Methodological aspects: Articles that followed both qualitative and quantitative methodology |
| **Databases** | Lens. Org, PubMed, Dimensions, Google Scholar |
| **search strategies** | ("Sex Worker*" OR "Prostitute*") AND ("Emergency Contraceptive*" OR "Emergency Contraceptive Pill*" OR "Morning After Pill*") |
| **Who do the screening and eligibility checking** | Authors independently/ independent reviewers. Resolving the discrepancies by discussion. |
| **Data items and definitions** | **01) Study Identification:** Author(s), year of publication, and title.<br>**02) Geographic Location:** Country or region where the study was conducted.<br>**03) Study Design:** Type of study (e.g., cross-sectional, qualitative, mixed-methods).<br>**04) Sample Size:** Total number of participants included in the study.<br>**05) Population Description:** Characteristics of the female sex worker population studied (e.g., street-based, entertainment workers, age group).<br>**06) Awareness of EC:** Awareness, understanding, misconceptions, and sources of information.<br>**07) Use of EC:** Prevalence of current or past use, frequency, and timing of use. |
|  | **08) Context of EC Use:** Situations leading to EC use, such as condom failure, stealthing, or sexual assault.<br>**09) Barriers to EC Use:** Reported challenges including stigma, cost, fear of side effects, limited access, or lack of awareness.<br>**10) Factors Associated with EC Use:** Socio-demographic or behavioural variables linked to increased or decreased EC use (e.g., age, education, marital status). |
| **Critical Appraisal of Individual Sources of Evidence** | In line with the objectives and established guidance for scoping reviews, a formal critical appraisal of the methodological quality of the included studies will not be conducted. The primary aim of this review is to map the extent, nature, and scope of existing literature on EC among FSWs, rather than to assess the risk of bias or inform outcome synthesis. As such, all eligible studies, regardless of design or methodological quality, will be included to ensure a comprehensive overview of available evidence.<br>Nonetheless, during the data charting process, attention will be given to study design and contextual limitations, which will be noted where relevant to support the interpretation of findings and highlight areas for future research. |
| **Analysis Method** | The final dataset of all included articles will be synthesized using a descriptive and thematic analysis approach. Quantitative data, such as prevalence rates, will be summarized using basic descriptive statistics. Qualitative findings will be organized and analyzed thematically under predefined categories aligned with the review objectives. |
| **Reporting Structure** | PRISMA-ScR guidelines |

## Annexure 02: Studies included for data charting

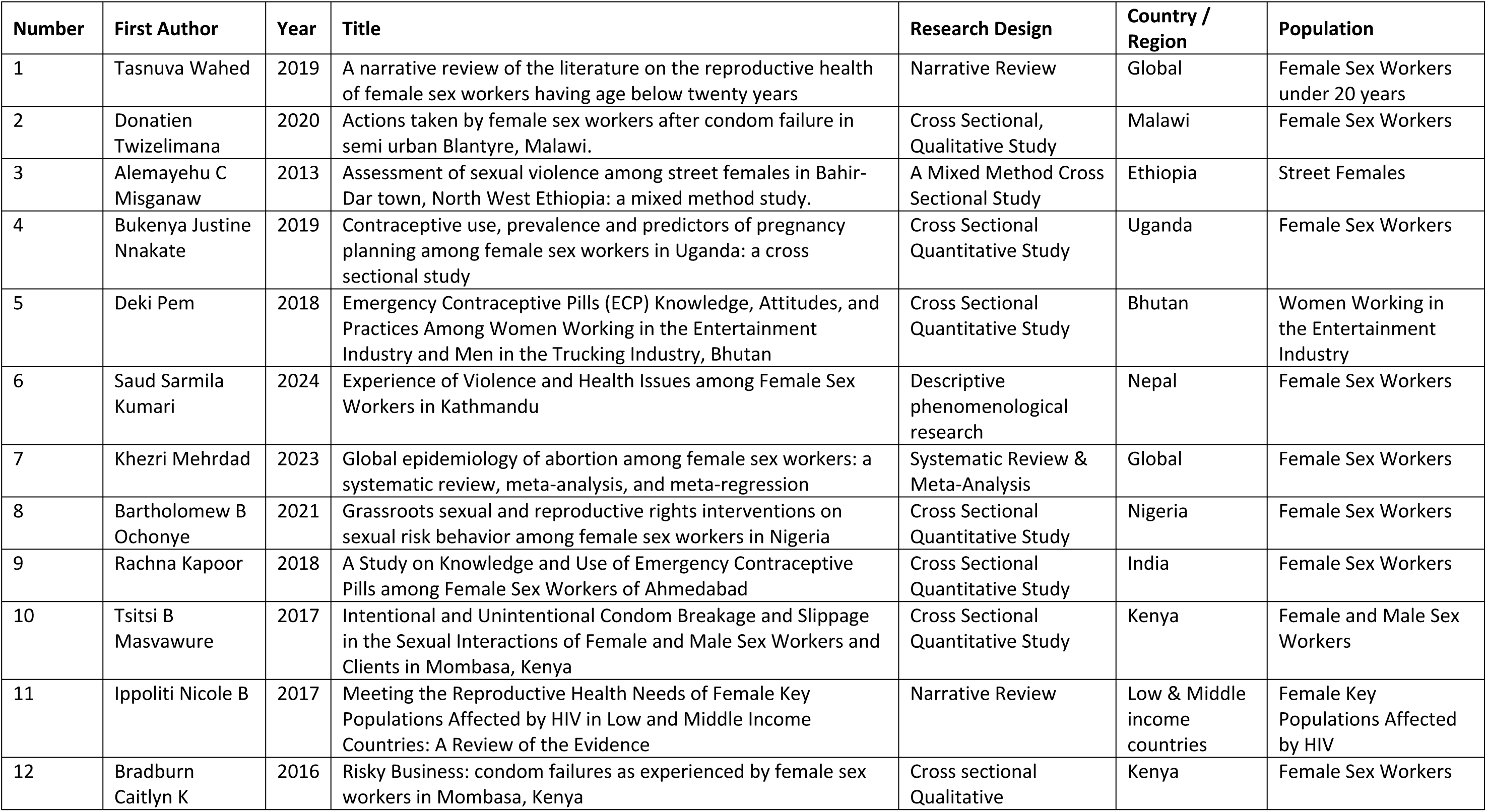
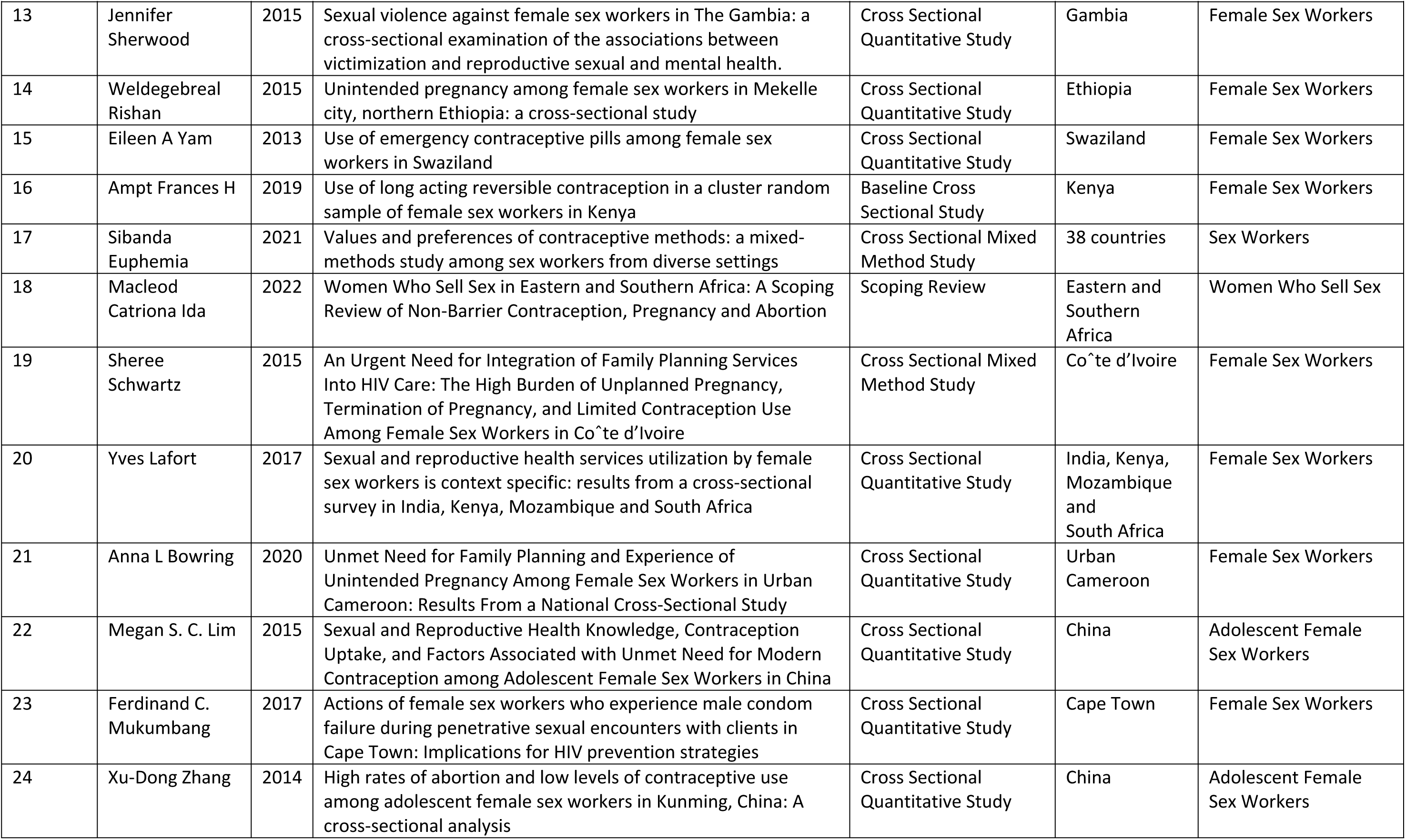

| Number | First Author | Year | Title | Research Design | Country / Region | Population |
| --- | --- | --- | --- | --- | --- | --- |
| 1 | Tasnuva Wahed | 2019 | A narrative review of the literature on the reproductive health of female sex workers having age below twenty years | Narrative Review | Global | Female Sex Workers under 20 years |
| 2 | Donatien Twizelimana | 2020 | Actions taken by female sex workers after condom failure in semi urban Blantyre, Malawi. | Cross Sectional, Qualitative Study | Malawi | Female Sex Workers |
| 3 | Alemayehu C Misganaw | 2013 | Assessment of sexual violence among street females in Bahir-Dar town, North West Ethiopia: a mixed method study. | A Mixed Method Cross Sectional Study | Ethiopia | Street Females |
| 4 | Bukenya Justine Nnakate | 2019 | Contraceptive use, prevalence and predictors of pregnancy planning among female sex workers in Uganda: a cross sectional study | Cross Sectional Quantitative Study | Uganda | Female Sex Workers |
| 5 | Deki Pem | 2018 | Emergency Contraceptive Pills (ECP) Knowledge, Attitudes, and Practices Among Women Working in the Entertainment Industry and Men in the Trucking Industry, Bhutan | Cross Sectional Quantitative Study | Bhutan | Women Working in the Entertainment Industry |
| 6 | Saud Sarmila Kumari | 2024 | Experience of Violence and Health Issues among Female Sex Workers in Kathmandu | Descriptive phenomenological research | Nepal | Female Sex Workers |
| 7 | Khezri Mehrdad | 2023 | Global epidemiology of abortion among female sex workers: a systematic review, meta-analysis, and meta-regression | Systematic Review & Meta-Analysis | Global | Female Sex Workers |
| 8 | Bartholomew B Ochonye | 2021 | Grassroots sexual and reproductive rights interventions on sexual risk behavior among female sex workers in Nigeria | Cross Sectional Quantitative Study | Nigeria | Female Sex Workers |
| 9 | Rachna Kapoor | 2018 | A Study on Knowledge and Use of Emergency Contraceptive Pills among Female Sex Workers of Ahmedabad | Cross Sectional Quantitative Study | India | Female Sex Workers |
| 10 | Tsitsi B Masvawure | 2017 | Intentional and Unintentional Condom Breakage and Slippage in the Sexual Interactions of Female and Male Sex Workers and Clients in Mombasa, Kenya | Cross Sectional Quantitative Study | Kenya | Female and Male Sex Workers |
| 11 | Ippoliti Nicole B | 2017 | Meeting the Reproductive Health Needs of Female Key Populations Affected by HIV in Low and Middle Income Countries: A Review of the Evidence | Narrative Review | Low & Middle income countries | Female Key Populations Affected by HIV |
| 12 | Bradburn Caitlyn K | 2016 | Risky Business: condom failures as experienced by female sex workers in Mombasa, Kenya | Cross sectional Qualitative | Kenya | Female Sex Workers |
| 13 | Jennifer Sherwood | 2015 | Sexual violence against female sex workers in The Gambia: a cross-sectional examination of the associations between victimization and reproductive sexual and mental health. | Cross Sectional Quantitative Study | Gambia | Female Sex Workers |
| 14 | Weldegebreal Rishan | 2015 | Unintended pregnancy among female sex workers in Mekelle city, northern Ethiopia: a cross-sectional study | Cross Sectional Quantitative Study | Ethiopia | Female Sex Workers |
| 15 | Eileen A Yam | 2013 | Use of emergency contraceptive pills among female sex workers in Swaziland | Cross Sectional Quantitative Study | Swaziland | Female Sex Workers |
| 16 | Ampt Frances H | 2019 | Use of long acting reversible contraception in a cluster random sample of female sex workers in Kenya | Baseline Cross Sectional Study | Kenya | Female Sex Workers |
| 17 | Sibanda Euphemia | 2021 | Values and preferences of contraceptive methods: a mixed-methods study among sex workers from diverse settings | Cross Sectional Mixed Method Study | 38 countries | Sex Workers |
| 18 | Macleod Catriona Ida | 2022 | Women Who Sell Sex in Eastern and Southern Africa: A Scoping Review of Non-Barrier Contraception, Pregnancy and Abortion | Scoping Review | Eastern and Southern Africa | Women Who Sell Sex |
| 19 | Sheree Schwartz | 2015 | An Urgent Need for Integration of Family Planning Services Into HIV Care: The High Burden of Unplanned Pregnancy, Termination of Pregnancy, and Limited Contraception Use Among Female Sex Workers in Co^te d'Ivoire | Cross Sectional Mixed Method Study | Co^te d'Ivoire | Female Sex Workers |
| 20 | Yves Lafort | 2017 | Sexual and reproductive health services utilization by female sex workers is context specific: results from a cross-sectional survey in India, Kenya, Mozambique and South Africa | Cross Sectional Quantitative Study | India, Kenya, Mozambique and South Africa | Female Sex Workers |
| 21 | Anna L Bowring | 2020 | Unmet Need for Family Planning and Experience of Unintended Pregnancy Among Female Sex Workers in Urban Cameroon: Results From a National Cross-Sectional Study | Cross Sectional Quantitative Study | Urban Cameroon | Female Sex Workers |
| 22 | Megan S. C. Lim | 2015 | Sexual and Reproductive Health Knowledge, Contraception Uptake, and Factors Associated with Unmet Need for Modern Contraception among Adolescent Female Sex Workers in China | Cross Sectional Quantitative Study | China | Adolescent Female Sex Workers |
| 23 | Ferdinand C. Mukumbang | 2017 | Actions of female sex workers who experience male condom failure during penetrative sexual encounters with clients in Cape Town: Implications for HIV prevention strategies | Cross Sectional Quantitative Study | Cape Town | Female Sex Workers |
| 24 | Xu-Dong Zhang | 2014 | High rates of abortion and low levels of contraceptive use among adolescent female sex workers in Kunming, China: A cross-sectional analysis | Cross Sectional Quantitative Study | China | Adolescent Female Sex Workers |

